# Poor metabolic health increases COVID-19-related mortality in the UK Biobank sample

**DOI:** 10.1101/2020.10.02.20205716

**Authors:** Filip Morys, Alain Dagher

## Abstract

Previous studies link obesity, components of metabolic health, such as hypertension or inflammation, to increased hospitalisations and death rates of patients with COVID-19. Here, in two overlapping samples of over 1,000 individuals from the UK Biobank we investigate whether metabolic health as measured by waist circumference, dyslipidaemia, hypertension, diabetes, and systemic inflammation is related to increased COVID-19 infection and mortality rates. Using logistic regression and controlling for confounding variables such as socioeconomic status, age, sex or ethnicity, we find that individuals with worse metabolic health (measured on average eleven years prior to 2020) have an increased risk for COVID-19-related death (adjusted odds ratio: 1.67). We also find that specific factors contributing to increased mortality are increased serum glucose levels, systolic blood pressure and waist circumference.

## Introduction

Since the beginning of the COVID-19 pandemic, mounting evidence supports an association between obesity and poor outcomes (1–12). The association holds for both obesity and for obesity-associated metabolic health – hypertension, diabetes, dyslipidaemia, and systemic inflammation (13). Similarly, obesity and excess adipose tissue have also been associated with higher risk of SARS-CoV-2 infection (10,14,15).

Previous studies, however, have tended to use small sample sizes, focus predominantly on the effects of body mass index (BMI) as a measure of obesity, or not account for confounding factors, such as ethnicity or socioeconomic status (14,16–18). Since ethnicity and socioeconomic status are themselves associated with obesity and metabolic health (19,20), they could confound interpretation of analyses in patients with COVID-19.

Here, we aim to present a comprehensive evaluation of obesity-associated metabolic risk factors that might be related to poor health outcomes in SARS-CoV-2 infected patients while controlling for confounding variables and limiting potential collider bias, which has previously resulted in incorrect epidemiological conclusions (21–23). We investigate whether metabolic health is related to higher chance for SARS-CoV-2 infection, but also COVID-19-related death.

## Materials and Methods

### Participants

In this study, we used the UK Biobank dataset – a large scale study with extensive phenotyping carried out in the United Kingdom (24). This study was performed under UK Biobank application ID 35605. SARS-CoV-2 real-time PCR test results in the UK Biobank dataset are derived from the Public Health England microbiology database Second Generation Surveillance System that is dynamically linked to the UK Biobank database (25). Here we only included individuals who were recorded as tested for SARS-CoV-2. We distinguished between two samples for two aims of our project: Sample 1 – a larger sample (n=12,659) of all individuals who were tested for SARS-CoV-2 between 16^th^ March 2020 and 24^th^ August 2020, to investigate the risk of COVID-19 infection and how it is related to metabolic health; and Sample 2, a subset of Sample 1 consisting of individuals who tested positive for COVID-19 (n=1,152). We also obtained data on mortality from COVID-19 for all the individuals included in our study population.

Sample characteristics can be found in Table 1. The discrepancies in number of patients who tested positive in both samples is due to outlier exclusions (see section: ‘Measures used in the study’). All participants signed written informed consent prior to participating in the UK Biobank study, which was approved by the North-West Multi-centre Research Ethics Committee (11/NW/0382). All UK Biobank actions are overseen by the UK Biobank Ethics Advisory Committee.

**Table 1.** Participants characteristics.

| Measure | Sample 1 (n=12,659) |  |  | Sample 2 (n=1,152) |  |  |
| --- | --- | --- | --- | --- | --- | --- |
|  | SARS-CoV-2 | SARS-CoV-2 | p- | SARS-CoV-2 | SARS-CoV-2 | p- |
|  | positive patients | negative patients | value | survivors | deceased | value |
|  | mean (SD) | mean (SD) |  | mean (SD) | mean (SD) |  |
| n | 1,138 | 11,521 |  | 972 | 180 |  |
| COVID-19 mortality rate |  | 1.42% |  |  | 15.63% |  |
| Age in years | 68 (8) | 70 (8) | <0.001 | 67 (9) | 74 (6) | <0.001 |
| Sex distribution | 47.36 % women | 52.32 % women | 0.002 | 49.07 % women | 36.67 % | 0.003 |
| Waist circumference (cm) | 93.38 (13.89) | 92.07 (13.69) | 0.003 | 92.55 (13.62) | 98.62 (14.48) | <0.001 |
| Serum HDL (mmol/l) | 1.36 (0.34) | 1.43 (0.38) | <0.001 | 1.37 (0.45) | 1.35 (0.32) | 0.446 |
| Serum TG (mmol/l) | 1.78 (1.04) | 1.77 (1.02) | 0.545 | 1.78 (1.05) | 1.88 (1.01) | 0.199 |
| HbA1c (mmol/mol) | 35.83 (4.51) | 35.76 (4.33) | 0.637 | 35.73 (4.62) | 36.99 (5.23) | **0.003** |
| Serum glucose (mmol/l) | 4.98 (0.64) | 5.00 (0.63) | 0.318 | 4.96 (0.63) | 5.19 (0.74) | **<0.001** |
| Serum C-reactive protein (mg/l) | 3.00 (5.23) | 2.84 (4.45) | 0.308 | 2.89 (5.14) | 3.98 (7.22) | 0.054 |
| Systolic blood pressure (mmHg) | 137.20 (18.59) | 138.55 (18.55) | **0.019** | 135.71 (18.15) | 145.76 (18.33) | **<0.001** |
| Diastolic blood pressure (mmHg) | 82.46 (10.25) | 82.24 (9.94) | 0.484 | 82.21 (10.31) | 84.03 (10.03) | **0.027** |
| Diabetes | 9.49 % | 7.83 % | 0.055 | 8.95 % | 15.56 % | **0.010** |
| Hypertension | 41.65 % | 42.32 % | 0.685 | 38.17 % | 62.22 % | **<0.001** |

SD – standard deviation, HDL – high density lipoprotein cholesterol, TG – triglycerides, HbA1C – haemoglobin A1c. P-values reflect significance of within samples differences calculated with t-tests in case of numerical variables and chi-squared tests in case of categorical variables.

### Measures used in the study

To investigate how metabolic health is related to SARS-CoV-2 infection and mortality rates, we used the following measures: waist circumference, serum triglyceride (TG), serum high density lipoprotein cholesterol (HDL), glycated haemoglobin (HbA1c), serum glucose (corrected for fasting times prior to blood drawing), serum C-reactive protein (26), previous diabetes diagnosis, resting systolic and diastolic blood pressure (mean of two measurements each), and hypertension diagnosis. In our analyses, we also controlled for age, sex, socioeconomic status (Townsend deprivation index (27), highest achieved educational qualifications, mean family income), smoking status, and ethnic background, the laboratory where COVID-19 testing was performed, and the origin of the sample used for COVID-19 test (e.g. nose, throat etc.). For Sample 2, we also used mortality data provided by the UK Biobank – COVID-19-related death was described using the ICD10 identifier U07.1. All variables related to metabolic health and all confounding variables were collected on average 11 years prior to COVID-19 tests.

Prior to the analyses, all numeric variables were z-scored, all serum level values were log-transformed, and we excluded outliers from the samples (2.2 interquartile range below 1st or above 3rd quartile). If participants were tested for SARS-CoV-2 more than once, they were considered positive if at least one test result was positive.

### Statistical analyses

The same analyses were performed for each of the two population samples. Data were analysed using R (v. 3.6.0). First, using confirmatory factor analysis in lavaan (v. 0.6-7), we estimated the fit of a latent variable ‘metabolic health’, which consisted of waist circumference, serum C-reactive protein levels, a latent variable ‘dyslipidaemia’ (serum HDL and TG levels; with loadings constrained to be equal between the two variables (28)), and two other latent variables, diabetes and hypertension. The latent variable ‘diabetes’ consisted of serum glucose and HbA1c levels, and diabetes diagnosis, while the latent variable ‘hypertension’ consisted of a blood pressure (systolic and diastolic blood pressure), and hypertension diagnosis (Figure 1). The model was estimated using robust maximum likelihood estimation and model’s fit was evaluated using common indices: comparative fit index (CFI), root mean square error of approximation (RMSEA), and standardized root mean square residual (SRMR). Acceptable fit was defined as CFI>0.9, RMSEA<0.1, and SRMR<0.08.

**Figure 1.**
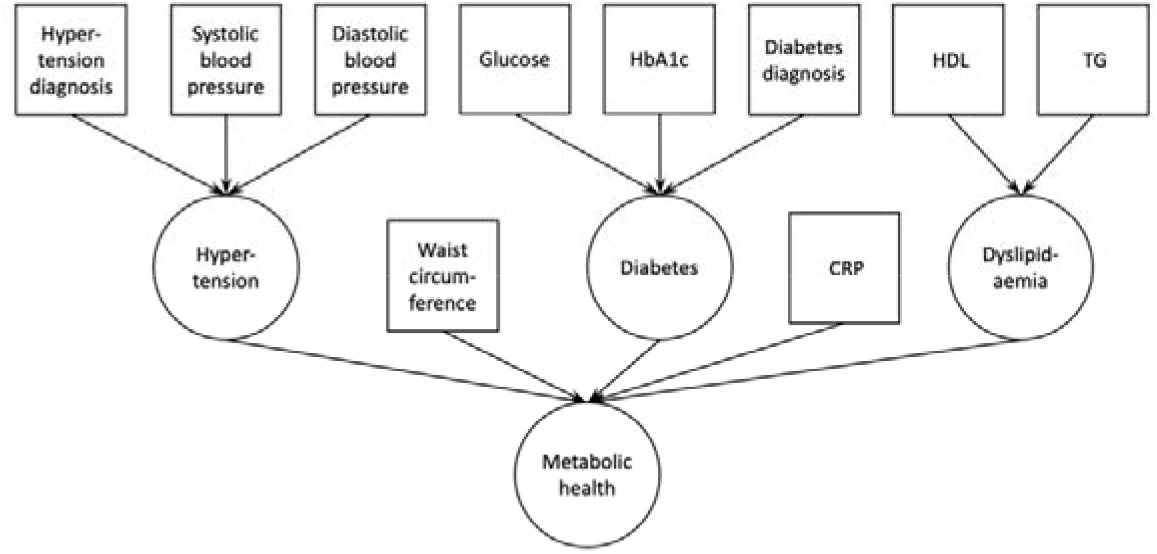
Confirmatory factor analysis model used to derive the latent variable ‘metabolic health. Squares indicate measured variables; circles indicate latent variables. HbA1c – glycated haemoglobin A1c; HDL – high density lipoprotein cholesterol; TG – triglycerides; CRP – C-reactive protein.

Next, we extracted components of the latent variable ‘metabolic health’ for each participant and entered them in a logistic regression. The outcome variable in logistic regression for Sample 1 was COVID-19 test result, while for Sample 2 the outcome variable was COVID-19-related death.

In the analyses we used a set of confounding variables to calculate adjusted odds ratio for testing positive for COVID-19: age, sex socioeconomic status, smoking status, ethnic background, test laboratory, and sample origin (6,29–31).

Finally, for Sample 2 we explored how individual factors contributed to the COVID-19-related mortality by using a logistic regression with individual components of metabolic health, instead of the latent variable ‘metabolic health’.

Overall, analyses using a latent variables approach allowed us to maximize information shared between different measured variables within the same metabolic domains, e.g. serum glucose levels, HbA1C levels and diabetes diagnosis for the latent variable diabetes. Furthermore, adding continuous measures in this study, such as glucose levels or blood pressure levels, instead of only using diabetes or hypertension diagnosis, enabled us to use more information that is available in the dataset and investigate in depth how continuous metabolic health measures are related to SARS-CoV-2 infection and COVID-19 mortality rates.

A script for the analysis of the data as well as the output of statistical software can be found at https://github.com/FilipMorys/COVID_MetS.

## Results

### Metabolic health and the risk of COVID-19 infections

In Sample 1, the confirmatory factor analysis provided an acceptable model fit (CFI=0.932, RMSEA=0.065, SRMR=0.040). In the logistic regression the relation between metabolic health and the chance of having a positive SARS-CoV-2 test did not reach our pre-set statistical significance threshold (p=0.060; odds ratio 1.10; 95% confidence intervals (CI): 0.996-1.216).

### Metabolic health is related to an increased COVID-19-related mortality

Among the individuals who tested positive, 180 people (16%) died from COVID-19, allowing us to investigate how metabolic health influences COVID-19-related mortality. Here, the confirmatory factor analysis provided an acceptable model fit (CFI=0.923, RMSEA=0.071, SRMR=0.049). Logistic regression showed that obesity-associated metabolic impairment was related to an increased mortality rate among COVID-19 positive individuals (p=0.0006) – adjusted odds ratio: 1.67 (95% CI: 1.25-2.23), pointing to a 67% increase for each unit increase on the metabolic health latent variable (Figure 2). Each unit increase on the metabolic health variable means increased serum glucose levels by 0.20 mmol/l, glycated haemoglobin A1c levels by 3.26 mmol/mol, C-reactive protein levels by 2.14 mg/l, triglyceride levels by 0.67 mmol/l, systolic blood pressure by 7.16 mmHg, diastolic blood pressure by 4.22 mmHg, waist circumference by 18.74 cm, and decreased HDL cholesterol levels by 0.25 mmol/l.

**Figure 2.**
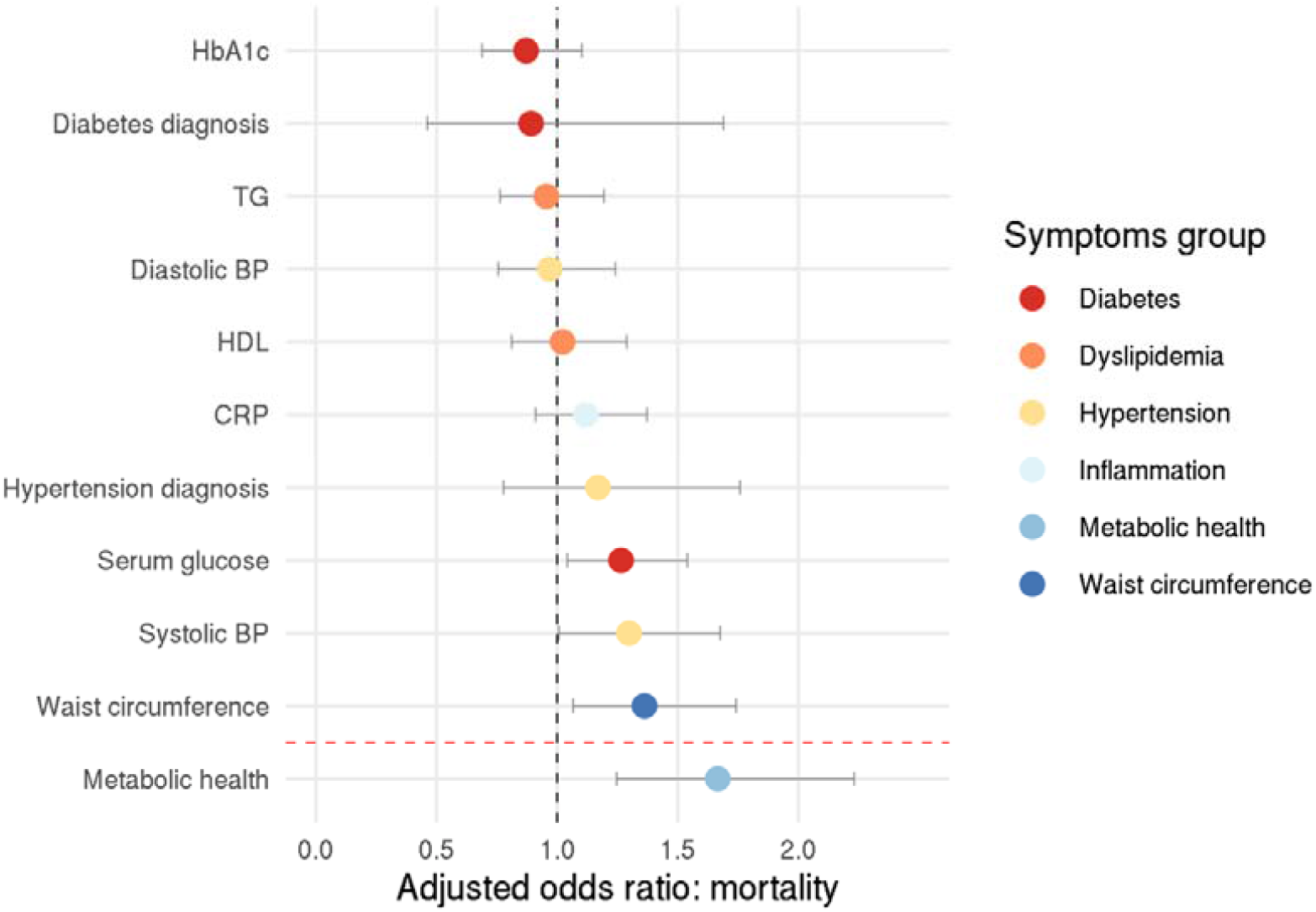
Odds ratio for COVID-19-related death. Circles represent adjusted odds ratios, error bars represent 95% confidence intervals.

Exploratory analysis with individual components of metabolic health revealed that higher systolic blood pressure, higher serum glucose, and higher waist circumference increased the chance of COVID-19-related mortality – the adjusted odds ratio were 1.30 (95% CI: 1.01-1.68), 1.26 (95% CI: 1.04-1.54), and 1.36 (95% CI: 1.07-1.74), respectively (systolic blood pressure: p=0.044, glucose levels: p=0.018, waist circumference: p=0.014; Figure 2).

## Discussion

We investigated whether obesity-associated poor metabolic health, here defined by increased waist circumference, increased TG levels, decreased HDL levels, systemic inflammation, increased glucose and HbA1C levels, increased systolic and diastolic blood pressure, and the presence of diabetes and hypertension, is a risk factor for COVID-19 infection and mortality. We were able to show that a one unit increase of the latent variable ‘metabolic health’ results in a 67% higher risk of death because of COVID-19. In contrast, the relation between metabolic health and the likelihood of test positivity was weaker, with an odds ratio of only 1.1 and a confidence interval that included the null effect. In sum, poor metabolic health contributes little to no risk of test positivity but substantially increases the odds of an adverse outcome. A strength of our analysis lies in the fact that the risk factor variables were measured prior to COVID-19 diagnosis and therefore were not influenced by the disease itself or possible treatment. Another is that we controlled for known shared risk factors between obesity and COVID-19 outcomes, such as socioeconomic status, ethnicity, and sex.

The results of our study are in line with previous reports linking obesity and individual components of metabolic health with poor COVID-19 outcomes and death (1,5,7,10,12,18,32). Bansal et al. review some of the physiological mechanisms that might mediate the relationship between obesity, related comorbidities and worse outcomes of COVID-19 (33). These include an enhanced expression of the angiotensin converting enzyme 2 (ACE2), diabetes-related microvascular dysfunction, increased expression of pro-inflammatory cytokines, or activation of the renin-angiotensin-aldosterone system related to hypertension (33–35). Recent reports especially highlight the role of interleukin-6 (IL-6) in the pathogenesis of COVID-19, but also the role of IL-6 inhibitors in COVID-19 therapy (36,37). This is important given that adiposity is associated with chronic systemic inflammation, which generally delays immune response to pathogens and can also lead to worse outcomes in COVID-19 patients (35,38–40). In general, our results suggest that a number of these mechanisms might also contribute to increased mortality rates from COVID-19.

Previous studies investigating COVID-19 in the UK Biobank used positive test results obtained between March 16^th^ and April 26^th^ as a proxy of severe COVID-19 (18,41). The rationale for this is that, during this time, only patients admitted to hospitals and with COVID-19-like symptoms were tested for SARS_CoV-2. Here, we decided to not use this approach for several reasons. First, it is possible that individuals with COVID-19-like symptoms for which they were admitted to a hospital but who did not have COVID-19 were only infected after being admitted. In those cases, positive tests would not reflect COVID-19 severity. Second, it is not possible to determine the exact reason for which inpatients were tested for SARS-CoV-2; positive test results might therefore not only reflect severity of COVID-19 disease, but also testing in anticipation of isolating patients admitted for other reasons. For example, patients having to undergo unrelated medical procedures might have undergone precautionary testing. We therefore recommend that studies that used this approach as a proxy for COVID-19 severity be interpreted with caution.

For the interpretation of our and similar results from the UK Biobank, it is important to note that the UK Biobank is not a sample representative of the entire UK population and therefore the findings might not be generalizable (42). Infection and mortality rates calculated here should not be used as an indicator of true prevalence and mortality rate in the general population. Furthermore, observational studies such as ours are prone to collider bias, which has already been identified in UK Biobank COVID-19 investigations (23). Collider bias occurs when the sample population is conditioned on a variable that correlates with the variables of interest. For example, during the time period of the current study, it is thought that health workers were more likely to be tested for SARS-CoV-2, which may have contributed to incorrect conclusion that cigarette smoking is protective, as health care workers have a lower incidence of smoking (23). Our study population includes individuals who were seen in a health care setting, who may therefore have a higher incidence of obesity and poorer metabolic health than the general population. However, it is difficult to see how this would account for the effect of obesity on death from COVID-19 among people who tested positive for the virus.

In addition, COVID-19 testing in the UK during the time period of this study was generally restricted to individuals with symptoms such as fever, cough, or loss of smell or taste – asymptomatic COVID-19 individuals were less likely to be tested. This might further increase the extent of collider bias in this and similar studies investigating the predictors of COVID-19 severity or mortality. Current strategies to account for such bias or measure the extent thereof rely on models and strong assumptions that might be incorrect. We therefore suggest that our findings be interpreted with caution. Nonetheless, our model did account for potential collider variables or confounds such as socioeconomic status, ethnicity and age.

Finally, in this study we only used limited data pertaining to COVID-19 diagnosis or mortality. The seriousness of illness, treatment administered, or actual cause of death were not investigated here.

In sum, we used the UK Biobank dataset to confirm that, in individuals who tested positive for COVID-19 in the early stages of the pandemic, metabolic health, and especially visceral adiposity, hypertension, and serum glucose levels, were associated with an increased risk of death.

## Data Availability

The data underlying the results presented in the study are available from the UK Biobank.

